# Prediction of Long COVID Based on Severity of Initial COVID-19 Infection: Differences in predictive feature sets between hospitalized versus non-hospitalized index infections

**DOI:** 10.1101/2023.01.16.23284634

**Authors:** Damien Socia, Dale Larie, Sol Feuerwerker, Gary An, Chase Cockrell

## Abstract

Long COVID is recognized as a significant consequence of SARS-COV2 infection. While the pathogenesis of Long COVID is still a subject of extensive investigation, there is considerable potential benefit in being able to predict which patients will develop Long COVID. We hypothesize that there would be distinct differences in the prediction of Long COVID based on the severity of the index infection, and use whether the index infection required hospitalization or not as a proxy for developing predictive models. We divide a large population of COVID patients drawn from the United States National Institutes of Health (NIH) National COVID Cohort Collaborative (N3C) Data Enclave Repository into two cohorts based on the severity of their initial COVID-19 illness and correspondingly trained two machine learning models: the Long COVID after Severe Disease Model (LCaSDM) and the Long COVID after Mild Disease Model (LCaMDM). The resulting models performed well on internal validation/testing, with a F1 score of 0.94 for the LCaSDM and 0.82 for the LCaMDM. There were distinct differences in the top 10 features used by each model, possibly reflecting the differences in type and amount of pathophysiological data between the hospitalized and non-hospitalized patients and/or reflecting different pathophysiological trajectories in the development of Long COVID. Of particular interest was the importance of Plant Hardiness Zone in the feature set for the LCaMDM, which may point to a role of climate and/or sunlight in the progression to Long COVID. Future work will involve a more detailed investigation of the potential role of climate and sunlight, as well as refinement of the predictive models as Long COVID becomes increasingly parsed into distinct clinical phenotypes.

## 1.0 Introduction

The development of long COVID carries significant morbidity for patients and a large financial burden to health systems. The diagnostic criteria for long COVID are quite broad; the diagnosis can incorporate numerous organ systems and the severity can range from mild to debilitating. This makes predicting which patients are at risk of developing the condition challenging. At the same time, the ability to predict the development of long COVID for a specific patient carries clinical promise and utility. Given the wide range of phenotypic presentations, however, we speculate that there is no unique patient signature at time of initial diagnosis of infection that can predict with absolute certainty if they will be affected by long COVID. We recognize that many features of long COVID resemble post-critical-illness syndrome, which in our clinical and scientific experience, is predicted by infectious disease severity. This provides a natural stratification for the patient population – those that were hospitalized when diagnosed with COVID-19 (and thus, we can infer they experienced a relatively severe illness), and those that were not hospitalized. In other words, those that experienced disease severe enough to require hospitalization and those that did not. Thus, we propose that the task of predicting long COVID must be divided into two distinct feature sets based on the above. Patients are split into two groups: Long COVID after Severe Disease (LCaSD) and Long COVID after Mild Disease (LCaMD). In this work, we utilized a suite of predictive variables ranging from personal health statistics to broader population demographics expecting that the severe disease model would be more informed by clinically measurable variables and the mild/moderate disease model would be more informed by patient histories and demographics. We posit that this unique approach yields a robust prediction tool.

## 2.0 Methods

The data source for this project was a subset of the National Institutes of Health (NIH) National COVID Cohort Collaborative (N3C) Data Enclave Repository [1]. Records of 58,000 of patients diagnosed with COVID-19 infection were available. 11,500 of these patients ultimately received a diagnosis of long COVID. The data repository contains billions of rows of relevant clinical patient data. As outlined above, we recognize the importance of separating the feature sets based on disease severity. Setting of care/hospitalization status was used a proxy for disease severity. Thus, we set out to develop two distinct prediction tools: the LCaSD and LCaMD models.

Our approach to constructing the prediction models can be divided into the following steps:

1. Aggregation and standardization (processing/cleaning) of patient-specific medical record fields
2. Selection and inclusion of potentially relevant historical, environmental and demographic features
3. Train machine-learning models on the developed datasets

### 2.1 Initial aggregation and standardization of data

The purpose of the initial aggregation/interpretation of patient medical records is to preemptively limit the number of putative features to be considered by the ML algorithm, given that the dataset is both extremely sparse and extremely broad. Lack of standardization in certain datasets (i.e., drug names, symptoms, devices, etc.) required data cleaning/pre-processing. Environmental and demographic variables were incorporated (at a coarse level), as these have been demonstrated to be predictive of a long COVID diagnosis [2,3]. XGBoost [4] was chosen as the ML library to build our models due to its efficiency in training and its ability to deal with missing data.

To aggregate and interpret patient-specific medical records, we considered phenotypic presentation at time of the initial COVID diagnosis, the setting of diagnosis (inpatient vs outpatient) prior diagnoses (comorbidities), symptomatic presentation post COVID-diagnosis, and drug/device history. Phenotypic presentation for the non-hospitalized cohort was defined by the values for heart rate, systolic and diastolic blood pressure, and oxygen saturation. For the hospitalized cohort, the above values were used along with measurements for serum creatinine, serum bilirubin, and platelet count; additionally, for the hospitalized cohort, the most deviant measure obtained during the hospitalization in which COVID is diagnosed for each vital sign/laboratory observable is used as input for the model.

### 2.2. Selection of potentially relevant features

As noted above, the pathophysiology of long COVID is still very unclear, and the empirical basis of the diagnosis of long COVID is in a state of evolution. Given that some authors [5] have identified instances of long COVID as a functional disorder without a clear biological signature, we sought to investigate whether there is a relationship between the presence of pre-existing functional disorder(s) and the risk of developing long COVID. We utilized the MIMIC-IV database [6] to identify the most common comorbidities for patients presenting with a set of six functional disorders (fibromyalgia, chronic pain, chronic fatigue, mast cell activation disorder, familial dysautonomia, and irritable bowel syndrome). We then classified the comorbidities into one of three categories: metabolic, psychiatric, or functional. For each patient, the number of prior diagnoses in each category was tabulated; thus, diagnostic history was compressed to three potential variables.

Additionally, membership into each of the predefined concept sets was used to further define patient diagnostic history. Symptomatic presentation post COVID was considered both in terms of the absolute number instances in which the patient sought medical care after a COVID diagnosis as well as specific symptoms which led the patient to seek care.

Symptoms were obtained from the available clinical note dataset. This exists in N3C as a post-processed dataset after a Natural Language Processing (NLP) algorithm procedure. Given the breadth of note data available, pre-processing was required to more precisely capture the symptoms of interest. First, only symptoms that were documented after the initial diagnosis of COVID were considered. We then removed all data rows which contained null values instead of clinical symptoms. Due to the prior NLP, clinical data points (symptoms or conditions) had been classified as “present,” “history of,” and “family history of.” We selected only the data points classified as “present” since our interest was symptom burden both during and after COVID-19 infection. We only selected data points that were NLP classified as referring to the patient as the subject. The NLP classified the certainty of data point presence as “positive,” “hypothetical,” “possible,” and “negated” (essentially representing a clinical review of systems schema often present in clinical notes). We selected only the data points classified as “positive.” Next, given the heterogeneity and redundancy in the reported symptom/concern field, a dictionary was created to match equivalent clinical terms (for example, “angina” = “ischemic chest pain” = “angina pectoris” and “cough” = “coughing” = “C/O - cough” = “does cough”). Finally, duplicate values that appeared for the same note date were dropped (only one complaint of “cough” was counted even if it was mentioned numerous times in the same note). The cleaned notes data was then aggregated on the patient level to obtain a total number of reported symptom occurrences after COVID diagnosis.

To develop a tractable drug history, we classified drugs from the available drug exposure dataset into 24 categories. For each drug category, the number of distinct drugs that fall within that category was tabulated. For device history, information related to respiratory assistance, harvested from the device exposure dataset, was used. Each hospitalized patient was then given a score, ranging from 0 to 5, based on the type of maximal respiratory assistance they required during hospitalization, with 0 indicating the patient received no support (ambient air) and 5 indicating that the patient received maximal support (mechanical ventilation).

Given the need for patient anonymity, the default location data available in N3C is limited to three-digit zip codes only. Thus, for the demographic and environmental features, we calculated the population-weighted-average for each variable per three-digit zip code using the populations of all five-digit zip codes that are contained within that given three-digit zip code. Populations were obtained from the 2010 census, at the level of ZIP Code Tabulation Area [7].

These variables include:

1. Plant-Hardiness Zone: a coarse metric representing climate [8]
2. Pandemic Mitigation (PM): a single metric approximating the relative pandemic mitigation. We derived this using the publicly available Google Community Mobility Reports [9], a dataset that provides a metric of social mobility for people who use Google Maps on their cellular telephone, defined by amount of time spent at a specific location categories (i.e., retail/recreation, grocery/pharmacy, parks, public transit, workplaces, and residential) relative to a six-week baseline of measurements that occurred prior to the implementation of societal PM strategies (colloquially referred to as “the lockdown”). We collapsed this metric to a single variable defined by calculating the daily sum of the relative amounts of time spent at workplaces; regions that observed more stringent PM protocols then spent relatively less time at work. After calculating the PM level for each region, the variable was normalized such that its values fall between 0 and 1.
3. 2018 Education Levels [10]
4. The 2013 Rural-Urban Continuum Codes [11]
5. Patient age, gender, and ethnicity

### 2.3. Predictive Model Training and Evaluation

The machine learning library used in this work was XGBoost. We first stratify the patient population into two subtypes – those that are hospitalized when diagnosed with COVID-19 and those that are not. Each of these sub-populations is then used to train a distinct prediction model using the XGBoost learning library. To determine the optimal features to use for each model, another model using all the features available was trained and tuned. The features were then sorted using the feature importance scores returned from XGBoost. New models were then iteratively trained, dropping the least important feature with each iteration. F1 scores were monitored to determine final feature set based on the highest F1. This was done for both the LCaSD and LCaMD prediction models. For hyperparameter tuning, six parameters were explored to find optimal performance; max depth of a single tree, the learning rate, the subsample ratio of the training instances sampled prior to growing trees, the subsample ratio of the features prior to growing trees, the subsample ratio of features for each level when a new depth level is reached in a tree, and finally the number of trees to include in the random forest. For each sub-population, using an 80/20 train/validate split of the training data and F1 as the scoring metric, a randomized grid search of the parameter space was performed for 1,000 iterations, with 5-fold cross validation, totaling 5,000 model fits for each sub-population. The parameters that scored the highest F1 score for each sub-population are the ones that are used in the final prediction models.

## 3.0 Results

Comparing the individual models, the F1 score for the LCaSD model (LCaSDM) was superior to the LCaMD model (LCaMDM): 0.94 vs 0.82 on available testing data. Examining the importance of features determined by each model, there were significantly fewer features used by the LCaSDM versus the LCaMDM (24 vs 54). Further, 17/24 of features used by the LCaSDM were objective health metrics (i.e., current disease status or clinically observable variable). Conversely, the data used by the LCaMDM were broader, incorporating patient histories, population-weighted demographics and environmental statistics via their anonymized three-digit zip code location. Top 10 features for each respective model are listed in Tables 1 and 2, respectively.

**Table 1:** Top 10 Features for Long COVID after Severe Disease Model (LCaSDM)

| LCaSDM Top 10 Features (by Relative Importance) |
| --- |
| Cold medicine (dextromethorphan, codeine, guaifenesin, benzonatate) |
| Obesity |
| Steroids (systemic) |
| Anticoagulation (tx and/or ppx) |
| Hospital visits post COVID diagnosis |
| Oxygen saturation |
| LOS |
| Inhalers |
| Metabolic syndrome (cluster) |
| Age |

**Table 2:** Top 10 Features for Long COVID after Mild Disease Model (LCaMDM)

| LCaMDM Top 10 Features (by Relative Importance) |
| --- |
| Vitamins/Electrolytes/Minerals |
| Visit ratio post COVID |
| Oxygen saturation |
| Visit ratio |
| Steroids (systemic) |
| Cardiomyopathy diagnosis |
| Hospital visits post COVID diagnosis |
| Plant hardiness zone |
| Inhalers |
| Education |

## 4.0 Discussion

The diagnosis of Long COVID remains a clinical dilemma, and an initial step in characterizing the pathophysiology of Long COVID involves being able to identify those patients who are risk of the disease. Risk prediction involves identifying sets of features about potential Long COVID patients, and we hypothesized that there would be distinct differences between those patients who were hospitalized for their index COVID infection and those who were not. Our work presented here demonstrates this difference in the important features used by each model, with a corresponding difference in the predictive value between them: F1 score of 0.94 for the LCaSDM versus 0.82 for the LCaMDM. We pose two potential (non-exclusive) interpretations for the significant difference in F1 scores between the models: 1) long COVID following severe illness bears a distinct data signature from that following mild/moderate illness; 2) higher resolution in clinical/laboratory data for hospitalized patients, along with the availability of more clinically objective data in this setting renders those features more predictive to the LCaSDM. Of note, one interesting predictive variable was plant-hardiness zone, chosen as an aggregate measure of climate for a region.

Future directions for this model involve utilizing five-digit zip codes combined with non-shifted COVID diagnosis dates to further investigate the link between climate/sunlight and long COVID and incorporating data on the likely SAR2-COV strains for given dates and locations. Future work will also involve subdividing the Long COVID patients into phenotypic clusters as has been recently reported/suggested by other investigators [12,13]; this will potentially provide insight into whether there are specific features or trajectories that can be correlated to clinical phenotypes.

## Conclusion

Utilizing a large national clinical database of patients with COVID-19 infection, we built two distinct models that predict the development of long COVID based on the severity of the initial COVID-19 illness. The resulting XGBoost models performed well, with F1 scores of 0.94 and 0.82 for the LCaSDM and LCaMDM, respectively. There were distinct differences between important predictive features depending on the severity of the index COVID infection, including an intriguing finding of the potential role of climate and/or sunlight on the risk of Long COVID. Future work will involve more detailed investigation of these potential climatic factors, as well as refining the prediction targets based on evolving recognition of Long COVID phenotypes.

## Data Availability

All data produced in this present study are available upon reasonable request to the authors

## Acknowledgements

The analyses described in this report were conducted with data or tools accessed through the NCATS N3C Data Enclave (https://covid.cd2h.org) and N3C Attribution & Publication Policy v 1.2-2020-08-25b supported by NCATS U24 TR002306 and NIH/NIGMS UO1EB025825. This research was possible because of the patients whose information is included within the data and the organizations (https://ncats.nih.gov/n3c/resources/data-contribution/data-transfer-agreement-signatories) and scientists who have contributed to the on-going development of this community resource [https://doi.org/10.1093/jamia/ocaa196].

## Authors Statement

CC conceived the project strategy and developed environmental and demographic inputs. DS and DL implemented the ML model, and performed data cleaning, feature selection, ML model evaluation. SF parsed and interpreted clinical data features, aided in model analysis and extensively edited the manuscript. GA assisted with project design and manuscript development.

